# The role of cytomegalovirus in prostate cancer incidence and mortality

**DOI:** 10.1101/2023.10.04.23296482

**Authors:** Johanna Classon, Abigail Britten, Kanar Alkass, Henrik Druid, Nicole Brenner, Tim Waterboer, Nicholas J Wareham, Effrossyni Gkrania-Klotsas, Jonas Frisén

## Abstract

Prostate cancer is one of the most common cancers in men with over 350 000 prostate cancer deaths reported worldwide every year. Current risk stratification models are insufficient to predict prostate cancer and prostate cancer death. New biomarkers are needed to identify those at increased risk of lethal prostate cancer. Cytomegalovirus (CMV) infection is common in the healthy prostate epithelium and promotes cell proliferation and viability in prostate cancer. Analyzing matched serum and tissue samples from post-mortem donors (n=41) and prostate cancer patients (n=40), we report that CMV seropositivity predicts high CMV abundance in prostate tissue. We studied if CMV seropositive men had increased prostate cancer incidence and cancer mortality in the European Prospective Investigation of Cancer (EPIC)–Norfolk population-based cohort study. CMV IgG serostatus was determined between 1993 and 2000 in 7,655 men aged 40-81 years, of which 57% were CMV seropositive. Study participants were followed for 18±6.0 years (mean±SD). We used Cox proportional hazard models, adjusted for age and potential confounders to estimate hazard ratios (HR) with 95% confidence intervals (CI). CMV serostatus was not associated with prostate cancer incidence (adjusted HR 1.03, 95% CI 0.89-1.19, p=0.687, 138,652 person-years). However, among prostate cancer patients, CMV seropositivity was associated with increased risk of prostate cancer-associated mortality (adjusted HR 2.26, CI 95% 1.02-4.99, p=0.044, 4639 person-years), with 25% of seropositive and 18% of seronegative prostate cancer patients dying from their disease during follow up. These results show that CMV seropositivity is associated with increased risk of prostate cancer death and suggest that CMV infection may contribute to prostate cancer lethality.

## INTRODUCTION

Prostate cancer is one of the most common cancers in men with over 350 000 prostate cancer deaths reported worldwide every year^1^. Whereas many prostate tumors are indolent, others can develop to metastatic disease, for which there is no cure. Few environmental factors have been linked to prostate cancer development, but the heritability of prostate cancer is 57%^2^, with more than 250 germline single nucleotide variants found to be associated with cancer incidence to date^3^. In contrast, only few germline events have been identified as risk factors for prostate cancer mortality, including carrying rare mutations in the gene *BRCA2*^4^, and potentially modifiable environmental factors may play a larger role in cancer progression. A probable relationship exists between smoking, obesity and advanced disease and risk for cancer progression, while physical activity might reduce this risk^5^. To date, tools that predict who will develop lethal prostate cancer are insufficient. For example, the commonly used Cancer of the Prostate Risk Assessment (CAPRA) score, which incorporates tumor features, s-PSA and age at diagnosis of localized prostate cancer, moderately predicts cancer-specific mortality (c-index 0.8)^6^. New biomarkers are essential to identify those at risk of lethal prostate cancer.

Cytomegalovirus (CMV) is a human herpesvirus that establishes chronic infection and globally 83% are seropositive to CMV^7^. However, seroprevalence differs markedly around the world, with western countries having much lower rates than countries in Africa and Asia^7^. CMV infection is present in a large fraction of normal prostate glands, prostate tumors and bone metastases^8,9^. In a case-control study, CMV serostatus was not associated with prostate cancer incidence, suggesting that CMV does not cause prostate cancer^10^. CMV seropositivity is, however, associated with increased all-cause mortality^11,12^. In the US NHANES III cohort, CMV seropositive men had increased cancer mortality^13^, but it is unknown if CMV serostatus is specifically associated with prostate cancer mortality. We recently reported that local CMV T-cell immunity in the prostate is associated with disease recurrence after radical prostatectomy in carriers of the common allele HLA-A*02-01^14^. Moreover, experimental CMV inhibition in models of prostate cancer can induce growth arrest and programmed cell death^9^.

CMV seropositivity likely underestimates the infected proportion of the population because CMV DNA is often detected along with low abundance of CMV responsive B-and/or T-cells in seronegative individuals^15–26^. These findings suggest that individuals are infected but have failed to mount a detectable antibody response. In cells of the hematopoietic lineage, CMV is more abundant in CMV seropositive persons, although CMV often is present but at lower levels in individuals without an antibody response^27–29^. Large interindividual variation exists in abundance of CMV infected epithelial cells in the prostate, ranging from 0-100%^9^, raising the question whether CMV serostatus could also be a marker of CMV abundance in the prostate.

Here we found that CMV was more abundant in benign prostates and prostate tumors in CMV seropositive compared to CMV seronegative men, demonstrating that CMV seropositivity serves as a marker for widespread CMV infection of the prostate. When we analyzed the large European Prospective Investigation of Cancer (EPIC)-Norfolk population-based cohort study, CMV serostatus was not associated with prostate cancer incidence. However, CMV seropositivity was associated with increased prostate cancer mortality in patients diagnosed with prostate cancer.

## METHODS

### Ethics Statement

Ethical permission for study of human samples was granted by the regional ethics committee of Sweden (2010/313-31/3). Prostate tissue and blood samples were collected from post-mortem donors between 2015 and 2020 through KI Donatum, Karolinska Institutet, Stockholm, Sweden, as described in^14^ and^9^; post-mortem cohort.

De-identified prostate cancer FFPE tissue from prostatectomy specimens with matched plasma (age span 50-68 years of age) was received from the Prostate Cancer Biorepository Network (PCBN) biobank; prostatectomy cohort. Ethical permission for biobank tissue collection was obtained from the local institutional review board at Memorial Sloan Kettering Cancer Center, New York, USA (Protocol #15-025).

The European Prospective Investigation of Cancer (EPIC)-Norfolk study was approved by the Norwich Local Research Ethics Committee (previously known as Norwich District Ethics Committee) (REC ref: 98CN01; 05/Q0101/191) and Waveney NHS Research Governance Committee (2005EC07L). All participants gave their informed written consent before entering the study. Permission to analyze EPIC-Norfolk data by Swedish researchers was granted by the Swedish Ethical Review Authority, ethical permit number 2021-00457 and its amendment 2022-00965-02.

### Analyses of CMV immunity in post-mortem and prostatectomy cohorts

We analyzed anti-CMV IgG titers in serum of post-mortem donors and in plasma of prostatectomy prostate cancer patients from PCBN using CMV IgG, CMIA assay (Architect, Abbott, Chicago, Illinois, USA). The cut-off value for a positive assay was 6.0 arbitrary units (AU) and measured values up to 250. Samples that had titers of 250 or higher were labeled ≥ 250. Four post-mortem donors were analyzed by CMV IgG chemiluminescence immunoassay on the LIAISON®XL Analyzer. Here >14 was considered positive (U/ml). We determined CMV IgG avidity using human anti-cytomegalovirus IgG ELISA Kit (ab108639, Abcam, Cambridge, UK) using low and high avidity controls (CMV IgG and IgG avidity EIA, DEIA465, Creative Diagnostics, New York, New York, USA). After sample incubation, wells were incubated with either 1X wash buffer or 6M urea in 1X wash buffer for 10 minutes. We calculated an index of CMV IgG avidity between non-urea and urea treated wells. High avidity: 0,80-1,00; Intermediate avidity: 0,65-0,79; Low avidity: <0,65.

Virscan data with Virscores were extracted from^14^, in which the analysis was further described, and compared with epithelial CMV-pp71 abundance as analyzed in ^9^. In brief, VirScan is a method to analyze antiviral IgG antibodies by using genetically engineered bacteriophages that display short peptides spanning viral genomes^30^. Prostate protein lysates were incubated with bacteriophages, IgG-bacteriophage complexes were captured using magnetic beads and sequenced. CMV VirScores, reflecting the number of CMV epitopes recognized by IgG antibodies in a sample, were determined as in^31^. The VirScan method is prone to false positive hits, and CMV VirScore of above five was here considered as CMV VirScore high.

### Analyses of prostate tissues from post-mortem donors and prostate cancer patients

Tissues were processed and analyzed as described in ^9^. Immunohistochemistry protocols and imaging are described in ^9^. This antibody was used: Cytomegalovirus pp71 (goat, 1:200, clone vC-20, sc-33323, polyclonal, Santa Cruz Biotechnology).

Histological evaluation of prostate cancer and inflammation was assessed in H&E stained FFPE prostate slides by a trained pathologist. Cells with lymphocyte morphology in large and small clusters or spread out in the stroma were classified as chronic inflammation. Acute inflammation with neutrophile infiltration was detected in three prostates in which chronic inflammation was also detected. An incidental prostate cancer (Gleason score 3+3) was detected in a CMV seronegative subject. A suspected incidental prostate cancer was detected in a CMV seropositive subject but was difficult to further assess due to post-mortal effects on the tissue. Two other individuals had been treated for prostate cancer with local radiation and chemotherapy respectively. Medical history was obtained from medical journals, next of kin, police reports and patient registry, all in accordance with ethical permit 2010/313-31/3.

In prostatectomy samples from PCBN (prostate cancer patients aged 50-68y), Gleason score and pathological T-stage were recorded by pathologists associated with PCBN. Gleason grade groups 1-5 were derived from Gleason scores according to the International Society of Urological Pathology.

### Statistical analyses of post-mortem and prostatectomy cohorts

All statistical analyses and plots for the cohort with post-mortem donors and the prostatectomy cohort were made in GraphPad prism 8.0 (GraphPad Software, San Diego, California, USA). Two-sided unpaired t-tests were performed on numerical continuous data with normal distribution. Groups with non-normal numerical continuous data was analyzed with Mann-Whitney test. Correlation analyses were performed with Pearson correlation analysis or Spearman correlation analysis when appropriate. Proportions between two groups was analyzed with Fisher’s exact test. Non-normal distributed data with more than two groups was compared with Kruskal-Wallis multiple comparisons. Receiver operator characteristic (ROC) curves were constructed and Area under the curve (AUC) calculated using % CMV-pp71 epithelial abundance in CMV IgG- and CMV IgG+ donors or patients. Patients with CMV IgG titers of ≥ 250 were all treated as having CMV IgG titer 250 in analyses.

### EPIC-Norfolk cohort characteristics

The EPIC-Norfolk study is a prospective population-based cohort in the UK that recruited study participants between 1993 and 1997, aged 40-79 years at time of inclusion. The cohort, with details on methods of data collection and follow up, has previously been described^12,32^. Participants subjected to CMV serology testing were randomly selected from the EPIC-Norfolk cohort^12,33^. End of follow-up was 2018 March 31^st^.

### Serum CMV IgG measurements in the EPIC-Norfolk cohort

In total, 7655 male study participants underwent CMV serology testing. CMV IgG serostatus was determined in serum from blood drawn on 1^st^ health check (1993-1998, n=5894) and/or 2^nd^ health check (1998-2000, n=3846). In blood collected at 1^st^ health check, serum CMV IgG measurements were performed using an indirect chemiluminescence immunoassay (Liaison, Diasorin, Saluggia, Italy). The CMV serology assessment procedure in blood samples collected at 1^st^ health check has previously been described in detail^12^. In blood collected at 2^nd^ health check, CMV serostatus was determined by assaying CMV epitopes, performed at the German Cancer Research Center, Heidelberg, Germany, as previously described^34^. In summary, antibodies against two or more antigens (CMV pp28, CMV pp52, CMV pp65, CMV pp150), with assay values above cut off values determined a priori, were considered as CMV seropositive.

### Variables collected at baseline in EPIC-Norfolk

Self-reported information on smoking, diabetes mellitus, alcohol usage, Townsend index^35^, education level, Body Mass Index, Waist-hip ratio, social class, ethnic origin, and marital status were collected at 1^st^ health check (1993-1998).

### Prostate cancer variables and endpoints in EPIC-Norfolk

Information on prevalent (present at determination of CMV serostatus) and incident prostate cancer (diagnosed after determination of CMV serostatus) was ascertained using ICD-10 code C61 in the UK Cancer Registry until 2016 March 31^st^, the latest available UK Cancer Registry data. Information on prostate cancer diagnosis, documented using ICD-10 codes 185, C61, was extracted from hospital admission and mortality data until 2018 March 31^st^. Hospital admissions data include all ICD codes recorded at admissions to hospital, independent of the reason. Prostate cancer diagnosis at hospital admissions is registered as date of admission and not backdated to the date the patient received his diagnosis. Death certificates were obtained for all participants who died before end of follow up at 2018 March 31^st^. Prostate cancer as underlying cause of death (185, C61) was used as the endpoint prostate cancer mortality.

Information on prostate cancer stage at diagnosis was documented in the UK cancer register for 284 prostate cancer patients, with 278 being incident prostate cancer cases, divided into general American Joint Committee on Cancer (AJCC) stage groups using TNM classifications. Patients were divided into categories 1-4 and 6, of which category 6 had insufficient information to produce a valid stage. Categories 1-4 were used, which correspond to cancer stage I-IV. Cancer was classified into localized prostate cancer (stage I-II; local growth; T2 or lower, N0, M0) and advanced prostate cancer (stage III-IV; tumor has invaded surrounding tissue and/or presence of lymph node metastases and/or presence of distant metastases; T3 or higher, or N1 or M1).

### Statistical analyses of EPIC-Norfolk

All analyses of EPIC-Norfolk data were executed in STATA SE 17 (Stata-Corp, College Station, Texas). All data available was included as specified below. No power calculations were conducted prior to this study. All tests of statistical significance were two-sided. P-value below 0.05 was considered statistically significant.

Characteristics for variables were summarized using mean and standard deviation (SD) for continuous variables and percentages for binary variables. Townsend index was used as a continuous variable, as was alcohol consumption in units per week. Age-adjusted linear regression analyses were performed to compare these variables between CMV seronegative and CMV seropositive males and prostate cancer patients. Binary variables: Prevalent Diabetes Mellitus (0/1), Ever smokers (0/1), Social class: employment and above (0/1), Education; A level and above (0/1; A level corresponds to 12 years of school education in the UK), Marital status; married or not married (0/1), Body Mass Index (≥ 30 or < 30 kg/m^2^), Waist-Hip ratio (≥ 0.90 or < 0.90; according to WHO definitions of high risk Waist-Hip ratio), Ethnic origin Caucasian or non-Caucasian (black Caribbean, black other, Indian, Pakistani, Chinese, Other). Binary variables were compared between CMV seronegative and CMV seropositive males using logistic regression adjusted for age. Data completeness was 97% or higher for all individual variables.

In time-to-event models, time from serostatus was determined using data from 1^st^ health check or 2^nd^ health check. These two health checks were two years apart. If a participant was CMV seronegative on both 1^st^ health check and 2^nd^ health check, time from CMV serostatus is determined as time from 1^st^ health check. If a participant was seronegative on 1^st^ health check and seropositive on 2^nd^ health check, time from CMV serostatus is determined as time from 2^nd^ health check. If a participant was seropositive on 1^st^ health check and seronegative on 2^nd^ health check, time from CMV serostatus is determined as time from 1^st^ health check (this pattern was observed for a rare number of study participants with CMV serostatus data from both 1^st^ health check and 2^nd^ health check; 47/1160: 4.1%). CMV IgG titers are stable over time^36^. As described in Table S2, CMV seroprevalence in matched age groups was lower in 2^nd^ health check compared to 1^st^ health check, implying that the sensitivity is lower, causing false negative CMV seronegative samples to occur.

Time to prostate cancer was determined as time to first prostate cancer registration, either in the prostate cancer registry or by registration upon hospital admission. Time from prostate cancer diagnosis to prostate cancer mortality was determined as time from first prostate cancer registration (Figure 2). Censored subjects in time-to-event analyses were either censored due to death from other causes than prostate cancer or were censored at end of study (2018, March 31^st^).

In time-to-event models, only patients with no missing data were analyzed. Time to event analyses were performed in study participants and patient with ≥ two years follow up excluding patients with prevalent prostate cancer. Time-to-event models were performed for all men from time of known serostatus to first prostate cancer registration or prostate cancer death and for prostate cancer patients from time of first registered prostate cancer diagnosis to prostate cancer death (Figure 2). Three time-to-event models were examined: 1) Unadjusted; 2) Adjusted for Age; 3) adjusted for Age, Smoking, Diabetes Mellitus, Body Mass Index, Waist-Hip Ratio, Education level and Townsend index. Assumptions of proportional hazards were examined by Schoenfeld residual statistical measures for each variable. If the assumption of proportional hazards was not fully justified for a variable, it was converted into a time-dependent variable in Cox proportional hazard models.

## RESULTS

### Relationship between CMV serostatus and CMV infection in the prostate

We recently reported that chronic CMV infection is common in the prostate using immunoblot, immunohistochemistry (IHC), quantitative PCR and in situ hybridization with the proportion of CMV infected cells in the prostate epithelium ranging from 0- 100%^9^. To assess the relationship between CMV infection in the prostate and CMV serostatus, we measured CMV IgG in serum of 41 post-mortem donors aged 19-89 in whom we quantified CMV abundance in the prostate.

In CMV seropositive post-mortem donors (24/41, 59%), the proportion of the prostate epithelial cells that was infected by CMV ranged from 2% to 100% with a mean of 58%, as assessed by the detection of the CMV protein pp71 by IHC (n=24, Figure 1A). It is well established that CMV infected individuals often are seronegative^15–26^, and prostates from the 17 CMV seronegative post-mortem donors were all CMV IHC positive, but epithelial abundance of CMV was significantly lower than in CMV seropositive post-mortem donors with a mean of 28% (p<0.0001, Figure 1A).

**Figure 1:**
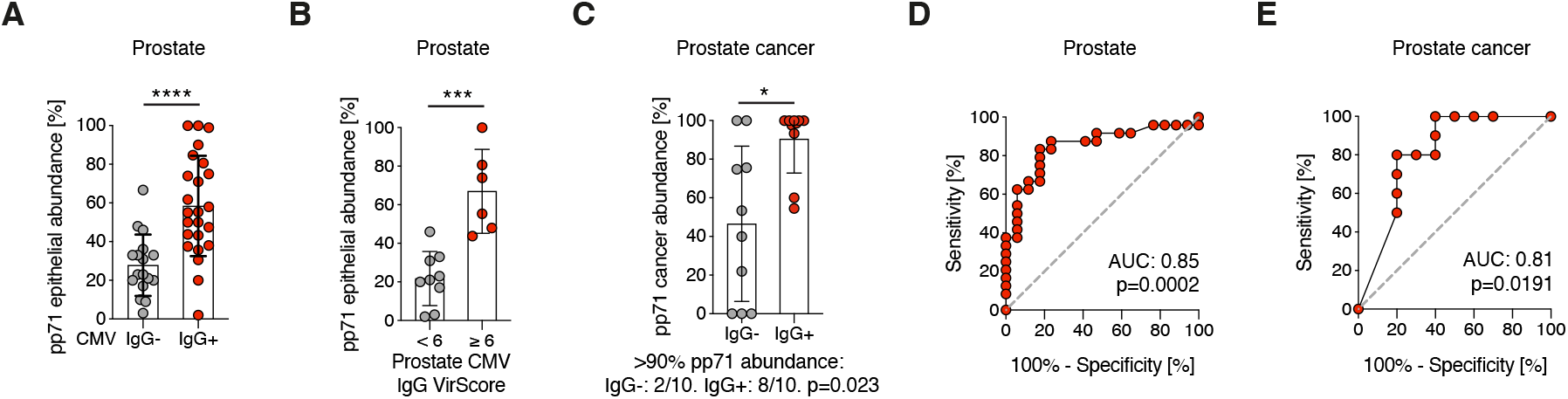
Relationship between CMV infection in the prostate and CMV serostatus. **A)** Quantification of percentage CMV-pp71 abundance in prostate areas with epithelial cells in prostate from post-mortem donors. Difference between CMV seronegative (IgG^-^; n=17) and CMV seropositive (IgG^+^; n=24) men was examined with un-paired two-tailed t-test. p<0.0001=****. Bar graph shows mean±SD. **B)** Abundance of CMV- pp71 (%) in prostates with high prostate CMV IgG VirScore (≥ 6) compared to low CMV IgG VirScore (< 6), n=15, analyzed with un-paired two-tailed t-test. P<0.001=***. Bar graph shows mean±SD. **C)** Abundance of CMV-pp71 (%) in tumors, determined with IHC, compared between CMV seronegative (IgG^-^; n=10) and CMV seropositive (IgG^+^; n=10) prostate cancer patients with Mann-Whitney test. Bar graph shows mean±SD. P<0.05=*. Analysis underneath graph shows Fisher’s exact test of fraction of samples with >90% cancer CMV abundance comparing CMV seropositive (2/10) and seronegative (8/10) patients. **D)** Receiver operator characteristic (ROC) curve for CMV-pp71 epithelial abundance (%) in CMV seropositive post-mortem donors in (A) (in red) compared to CMV seronegative post-mortem donors in (A) (in grey). AUC = Area under the curve. **E)** ROC curve for CMV-pp71 cancer abundance in CMV seropositive prostate cancer patients in (C) (in red) compared to CMV seronegative prostate cancer patients in (C) (in grey). AUC = Area under the curve.

We also found a positive association between a high CMV IgG VirScore in prostate tissue^14,30^, reflecting presence and diversity of CMV IgG antibodies, with CMV-pp71 epithelial abundance (n=15, p=0.003) (Figure 1B), further supporting an association between CMV immunity and epithelial CMV infection. Prostates of CMV seropositive donors did not have higher prevalence of inflammatory infiltrates with lymphocytes than in CMV seronegative donors (Figure S1A). Neither was presence of inflammatory infiltrates associated with CMV abundance in CMV seronegative nor CMV seropositive donors (Figure S1B), ruling out general chronic inflammation as a mediator.

A more pronounced association between CMV serostatus and prostate CMV abundance, determined with CMV-pp71 IHC, was observed in prostate cancer patients who underwent prostatectomy (Prostatectomy cohort). First, 3/10 of prostatectomy samples from CMV seronegative prostate cancer patients had CMV negative tumors and CMV negative benign epithelium (Figure 1C). Second, CMV was commonly found throughout the whole tumor in CMV seropositive patients (Figure 1C, n=10) whereas CMV was less abundant in the tumor in CMV seronegative patients (IgG+ vs IgG-, >90% CMV^+^: p=0.023, n=10 per group, Figure 1C). The median CMV tumor abundance was 99% in CMV seropositive patients (n=10) and 47% in CMV seronegative patients (n=10) (p=0.016; Figure 1C).

With higher age, titers of anti-CMV IgG antibodies increase, perhaps due to subclinical reactivation events^28,37^. Prostate CMV abundance is positively correlated with age^9^, but when we analyzed CMV seronegative and CMV seropositive donors separately, CMV abundance only increased with age in CMV seronegative donors (CMV IgG^-^: r=0.72, p=0.001; CMV IgG^+^: r=0.25, p=0.24; Figure S2A). Furthermore, serum CMV IgG titer did not correlate with prostate CMV abundance in CMV seropositive donors (r=0.03, p=0.90, Figure S2B) nor CMV abundance in prostate tumors in CMV seropositive prostate cancer patients (r=0.28, p=0.44, Figure S2C). High avidity of serum CMV IgG antibodies did not further discriminate CMV abundance (Figure S2D). In conclusion, measures of systemic CMV humoral immunity quality (IgG avidity) and quantity (IgG titer) could not be used to discriminate CMV abundance.

We asked how well CMV serostatus distinguished abundance of CMV in prostate epithelium. Receiver operator characteristic (ROC) analyses showed that CMV seropositivity identifies prostates with higher CMV abundance in post-mortem donors (Area under the curve (AUC): 0.85 (CI 95% 0.73-0.97), p=0.0002) (Figure 1D) and in primary prostate tumors (AUC: 0.81 (CI 95% 0.61-1.00), p=0.0191) (Figure 1E).

In summary, CMV seropositive men have higher abundance of CMV infected prostate cells than CMV seronegative men. Hence, CMV serostatus is a marker for CMV abundance in benign and malignant prostate tissue.

### EPIC-Norfolk male CMV cohort characteristics

Next, we asked if CMV infection was associated with prostate cancer incidence or mortality. In order to do so, a large number of prostate specimens or blood samples are required. Since there is often CMV heterogeneity within a prostate, accurate assessment of CMV would entail analyzing large areas, which is difficult to do to in large scale. In contrast, CMV serostatus is analyzed with scalable assays in blood samples.

We used CMV serostatus to examine if CMV was associated with prostate cancer incidence and mortality (Figure 2A-B). In EPIC-Norfolk, a total of 7655 male participants aged 40-79 years at study recruitment in 1993-1997 were examined for CMV serostatus^12,33^. CMV serostatus was determined in serum from blood drawn 1993-1998 (n=5894) and/or 1998-2000 (n=3846) in males aged 40-81 years. The study participants were followed for 18±6.0 years (mean±SD) from blood draw until the end of follow up due to death or end-of-study. Of the study participants, 57% were CMV seropositive on at least one blood test (Table S1, Figure 2). As expected, since CMV seropositivity increases with age^38^ (Table S2), CMV seronegative participants were on average 2.9 years younger than CMV seropositive participants (58.6±9.1 vs 61.5±9.2 years; mean±SD) (Table S1). In age-adjusted analyses, CMV seropositive men were more likely to be non-white, smokers, have high BMI, be married, have lower level of education and had addresses in areas of material deprivation, as measured by Townsend index score (Table S1).

**Figure 2:**
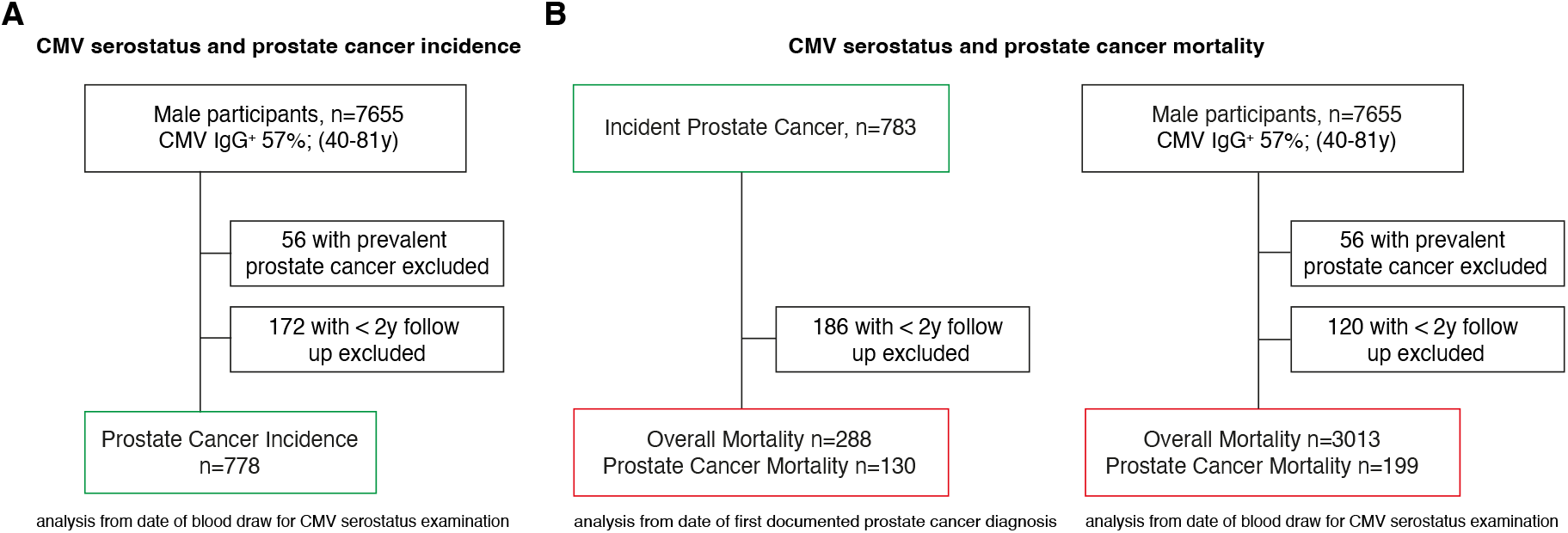
Flow charts of EPIC-Norfolk CMV and prostate cancer study. **A)** Study design of the EPIC-Norfolk cohort in which CMV serostatus (CMV IgG^-^ vs CMV IgG^+^) and risk of prostate cancer (prostate cancer incidence) was examined. Patients with prevalent prostate cancer and those with less than two years follow up time from CMV serostatus determination to a documented prostate cancer diagnosis were excluded. **B)** Study design of the EPIC-Norfolk cohort in which CMV serostatus (CMV IgG^-^ vs CMV IgG^+^) and risk of overall mortality and prostate cancer mortality was examined. All men in the cohort, excluding patients with prevalent prostate cancer and those with less than two years follow up time from CMV serostatus determination to death were analyzed. Patients with incident prostate cancer with more than two years follow up were examined in independent analyses. Green boxes indicate prostate cancer cases. Red boxes indicate mortality as endpoint in analyses.

Prostate cancer diagnoses were identified through the UK cancer registry (n=759) and/or by hospital admissions (n=693), identifying a total of 839 prostate cancer patients (Figure S3). Most patients were diagnosed with prostate cancer after CMV serostatus was defined (incident cases, n=783, 93% of all cases) and 219 died from prostate cancer during the study period. Incident prostate cancers were registered 11.9±6.0 years after CMV serostatus analysis.

Twenty prostate cancer patients had their cancer diagnoses registered at the same time as death from the disease, which is likely due to delayed registration of diagnosis. In addition, 11/205 (5.4%) of patients with localized prostate cancer stage at diagnosis in the Cancer Registry died within two years, further substantiating that this error occurs, as the 2-year survival of localized prostate cancer is nearly 100%. To account for this bias, all time to event analyses were performed in study participants and patient with ≥ two years follow up.

### CMV serostatus and prostate cancer incidence

In total, 483 (61.7%) of the men diagnosed with prostate cancer during follow-up were CMV IgG^+^ at the time of blood draw. CMV seropositive men were on average 2.5 years older (mean 76.1± SD 7.9) than CMV seronegative men (mean 73.6± SD 7.8) when their incident prostate cancer diagnoses were registered. CMV seropositivity was not associated with increased risk of prostate cancer incidence during follow up in Cox proportional hazard models adjusted for age (HR 0.99, 95% CI 0.85-1.14, p=0.848) or smoking, diabetes mellitus, body mass index, waist-hip ratio, education level and Townsend index in addition to age (HR 1.03, 95% CI 0.89-1.19, p=0.687) (Table 1). Estimated crude incidence rate ratios (IRR) did not show an association between CMV and prostate cancer incidence (IRR 1.14 95% CI 0.99-1.32, p=0.068, 138,652 person-years) (Table S3). In addition, prostate cancer patients aged 50-68 years who underwent prostatectomy (prostatectomy cohort) were not largely CMV seropositive (CMV IgG-: 19/40, CMV IgG+: 21/40, Figure S4A). In conclusion, we find no evidence of a general association between CMV and prostate cancer incidence.

**Table 1:** CMV seropositivity and prostate cancer incidence. *Adjusted for Age, Townsend index, Diabetes Mellitus, Body Mass Index, Waist-Hip Ratio, Education level and Smoking. Cox proportional hazard models. CI: Confidence Interval.

|  | **Hazard ratio (CI 95%)** | **p-value** |  |
| --- | --- | --- | --- |
| Model 1 (Un-adjusted) | 1.17 (1.01-1.34) | 0.035 | n=7427, 778 events |
| Model 2 (age-adjusted) | 0.99 (0.85-1.14) | 0.848 | n=7427, 778 events |
| Model 3 (fully adjusted)* | 1.03 (0.89-1.19) | 0.687 | n=7324, 766 events |

### CMV and prostate cancer characteristics

We next asked if CMV serostatus is associated with known prognostic prostate cancer features. CMV seropositivity was not more prevalent in patients with advanced compared to localized prostate cancer in EPIC-Norfolk at time of diagnosis (n=278, age-adjusted p=0.487, advanced prostate cancer CMV seronegative: 31% and CMV seropositive: 27%; localized prostate cancer CMV seronegative: 69% and CMV seropositive: 73%, Table S4). In the prostatectomy cohort (n=40), prostate cancer patients had T-stage pT2 or pT3 tumors with varying Gleason scores. CMV seropositivity was not associated with T-stage (p=0.205; Fisher’s exact test; Figure S4B) nor Gleason grade group (p=0.078; Fisher’s exact test; Figure S4C). CMV abundance in primary prostate tumors (n=20) was not significantly associated with Gleason score (grade group) or T-stage (Figure S4D-E, prostatectomy cohort). In summary, CMV serostatus was not associated with the known prostate cancer prognostic risk factors advanced prostate cancer stage, Gleason score or T-stage at diagnosis, but appears to be an independent parameter.

### CMV serostatus and prostate cancer mortality

Examining all men in the EPIC-Norfolk cohort, an association between CMV serostatus and prostate cancer mortality was not apparent (adjusted HR 1.24, CI 95% 0.92-1.67, p=0.166, n=7375, Table 2) but an effect of CMV seropositivity on all-cause mortality adjusted for age and potential confounders was found (adjusted HR 1.10, CI 95% 1.02-1.18, p=0.019, n=7375, Table 2). Crude incidence rate ratios for all mortality analyses are reported in supplemental Table 7.

**Table 2:** CMV seropositivity, prostate cancer mortality and all-cause mortality. Prostate cancer patients are defined as incident cases, i.e. that their diagnoses are registered after CMV serostatus examination. CMV seropositive patients are compared to CMV seronegative patients. *Adjusted for Age, Townsend index, Diabetes Mellitus, Body Mass Index, Waist-Hip Ratio, Education level and Smoking. Cox proportional hazard models. CI: Confidence Interval.

|  | Hazard ratio (CI 95%) | p-value |  |
| --- | --- | --- | --- |
| **All-cause mortality in patients with prostate cancer** |  |  |  |
| Model 1 (Un-adjusted) | 1.35 (1.06-1.72) | 0.014 | N=597, 288 events |
| Model 2 (age-adjusted) | 1.21 (0.95-1.54) | 0.121 | N=597, 288 events |
| Model 3 (fully adjusted)* | 1.17 (0.92-1.50) | 0.206 | N=588, 281 events |
| **Prostate cancer mortality in patients with prostate cancer** |  |  |  |
| Model 1 (Un-adjusted) | 2.47 (1.12-5.43) | 0.024 | N=597, 130 events |
| Model 2 (age-adjusted) | 2.32 (1.06-5.13) | 0.036 | N=597, 130 events |
| Model 3 (fully adjusted)* | 2.26 (1.02-4.99) | 0.044 | N=588, 126 events |
| **All-cause mortality in all men** |  |  |  |
| Model 1 (Un-adjusted) | 1.45 (1.34-1.56) | <0.001 | N=7479, 3013 events |
| Model 2 (age-adjusted) | 1.11 (1.03-1.20) | 0.006 | N=7479, 3013 events |
| Model 3 (fully adjusted)* | 1.10 (1.02-1.18) | 0.019 | N=7375, 2960 events |
| **Prostate cancer mortality in all men** |  |  |  |
| Model 1 (Un-adjusted) | 1.61 (1.20-2.17) | 0.001 | N=7479, 199 events |
| Model 2 (age-adjusted) | 1.24 (0.92-1.67) | 0.155 | N=7479, 199 events |
| Model 3 (fully adjusted)* | 1.24 (0.92-1.67) | 0.164 | N=7375, 193 events |

We next examined if CMV serostatus was associated with all-cause and prostate cancer mortality specifically among patients diagnosed with prostate cancer. Of incident prostate cancer patients, CMV seronegative patients were followed for 7.9±4.3 years (mean±SD) and CMV seropositive patients were followed for 7.6±4.4 years (mean±SD) after prostate cancer registration. CMV seropositive prostate cancer patients did not display increased all-cause mortality compared to CMV seronegative prostate cancer patients, as examined in Cox proportional hazard models adjusted for age and other potential confounders (fully adjusted HR 1.17, CI 95% 0.92-1.50, p=0.206, n=588, Table 2).

During follow up, 24.7% of CMV seropositive patients and 17.6% of CMV seronegative prostate cancer patients died from their disease. CMV seropositivity was associated with higher risk of prostate cancer mortality in unadjusted (HR 2.47, CI 95% 1.12-5.43, p=0.024, n=597) and age-adjusted (HR 2.32, CI 95% 1.06-5.13, p=0.036, n=597) Cox proportional hazard models. The association persisted after adjusting for the potential confounders smoking, diabetes mellitus, body mass index, waist-hip ratio, education level and Townsend index in addition to age (HR 2.26, CI 95% 1.02-4.99, p=0.044, n=588) (Table 2).

We conclude that CMV seropositivity, which correlated with higher abundance of CMV in prostate epithelial cells, was associated with increased cancer mortality in prostate cancer patients.

## DISCUSSION

We report that CMV infection is more widespread in epithelial cells in the healthy prostate and in prostate cancer in CMV seropositive compared to seronegative men, establishing CMV serostatus as a marker for CMV abundance in the prostate. In a large prospective cohort, CMV serostatus was not associated with prostate cancer incidence, corroborating previous research that found no association between DNA virus immunity and risk of prostate cancer^10,39,40^. These findings imply that CMV is not an important cause of primary prostate cancer. We, however, report a positive association between CMV seropositivity and prostate cancer mortality in prostate cancer patients, independent of potential confounders such as obesity, smoking and low socioeconomic status.

Many men who were CMV seronegative still had CMV infected prostates. This is not surprising, as it is well established that CMV serostatus substantially underestimates the proportion of individuals who are infected by CMV^15–22^. Thus, we are not exclusively comparing groups of individuals who are CMV infected or not, but who are infected to varying degrees. Importantly, this raises the possibility that CMV potentially also may affect prostate cancer mortality in the CMV seronegative group, but to a lesser extent, and that the increased mortality after prostate cancer diagnosis in seropositive prostate cancer patients may underestimate the true effect of CMV. Interestingly, CMV IgG titer in blood could not be used to distinguish prostates with highest CMV abundance. Factors such as age at infection and CMV strain variability may be more important.

This study has several strengths, including long follow-up time and high data completeness. Furthermore, no study participants were lost during follow up other than due to death prior to study completion. This study also has limitations. Over 99% of study participants were of Caucasian ethnicity, all living in the UK. It is therefore difficult to generalize the study results to other populations such as African American men in USA or men in east Asia, two groups that display increased risk of prostate cancer mortality and have high CMV seroprevalence^1,7,38,41^. Although excluding participants with less than two years follow up potentially reduced effects of bias introduced by late registration of prostate cancer diagnoses, this strategy may potentially exclude a small fraction of prostate cancer patients diagnosed with *de novo* metastatic prostate cancer with poor outcome.

CMV seropositivity may be a marker of an altered immune system or of a yet unidentified confounder that could explain the observed associations. Additionally, CMV seropositivity could perhaps select for an earlier death from non-cancer causes and result in selective survival bias, as CMV seropositivity is associated with all-cause mortality and particularly mortality from cardiovascular disease^11,12^. However, we expect that any possible bias by selective survival is small and would not account for our observations. A main cause of death among prostate cancer patients is cardiovascular disease, and may therefore be a competing risk of death, particularly among CMV seropositive men^42^. An association between CMV serostatus and prostate cancer mortality is not as evident when analyzing all men in our cohort from age of CMV serology, which may relate to the lack of increased incidence and competing hazards.

The proportion of prostate cancer patients in EPIC-Norfolk diagnosed via s-PSA testing or by showing clinical signs is unknown but could impact study results. CMV seronegative men may be more likely to enroll in s-PSA testing as they in general have higher socioeconomic status than CMV seropositive men (Table S1), which could increase the number of indolent prostate cancer cases in CMV seronegative men. However, adjusting for education level, Townsend index and smoking, among other factors, did not reduce the effect measure of CMV’s association with prostate cancer mortality (Table 2). Furthermore, the proportion of patients with advanced cancer disease at diagnosis in EPIC-Norfolk was not dependent on CMV serostatus (Table S3), which suggests that confounding due to different diagnostic intensity is improbable. Rather, trends in EPIC-Norfolk and prostatectomy cohorts suggest that CMV seronegative patients may perhaps be diagnosed with prostate cancer with higher T-stage and Gleason scores than CMV seropositive patients (Table S4, Figure S4B-C). Previous research has shown that there is no association between CMV serostatus and Gleason score^10^. The limited information on prostate cancer features and low patient number in our study impede further exploration.

CMV may drive prostate cancer progression and metastatic spread independent of cellular differentiation (represented by Gleason score) and local invasion (represented by T-stage), perhaps through early seeding. This is in contrast to prostate cancer associated bacteria that may be associated with Gleason score^43^ and characteristics of advanced disease^44,45^. This could be due to direct and indirect effects of CMV, For example, CMV-directed T-cells can skew immune responses^46^ and potentially be pro-metastatic. In prostate cancer models, CMV directly promotes cell viability and proliferation^9^, two crucial features of malignant cells that allow tumors to accumulate alterations, perhaps induced by CMV, that result in metastatic disease.

It will be important to validate the potential of CMV seropositivity as a risk factor for prostate cancer mortality in clinical cohorts, to further examine associations with clinical factors, including treatment modality, s-PSA and family history of prostate cancer. The potential added value of a CMV based assay, such as CMV serology, in prostate cancer prognosis models will be essential to define but was not possible to determine in this study. CMV promotes androgen signaling in prostate cancer and can modulate cellular processes that are associated with resistance to therapies for advanced prostate cancer^9^. It will be important to define if CMV is a predictive marker for the efficiency of anti-androgen therapies. Drugs that target CMV resulted in inhibited growth and induced cancer cell death in CMV infected prostate cancer models^9^. CMV serostatus has the potential to guide treatment choices and direct patient treatment in clinical trials of anti-viral therapies such as maribavir and letermovir^9^, with the hope that these, or other CMV targeted drugs, will prolong survival of prostate cancer patients.

## Data Availability

All data produced in the present work are contained in the manuscript.

## ACKNOWLEDGEMENTS

All authors contributed to the study conception, study design, analysis, and interpretation of data. J.C and J.F wrote the manuscript with input from all authors. We thank the MRC Epidemiology Unit for access to data in the EPIC-Norfolk cohort. We thank the Karolinska University Hospital Clinical Microbiology Laboratory where CMV serology assays were performed for post-mortem and prostatectomy cohorts. We thank Professor Hans-Olov Adami and Lars Björnebo for valuable input on the manuscript. J.F and E.GK had full access to all the data in the study and takes responsibility for the integrity of the data and the accuracy of the data analysis.

