## Supplemental figures and tables for "The role of cytomegalovirus in prostate cancer incidence and mortality"

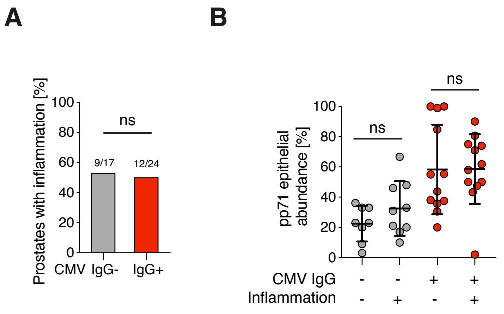


**Supplemental Figure 1: CMV abundance is not associated with chronic inflammation in prostate**

**A)** Inflammation was histologically examined in in H&E prostate sections. Lymphocyte infiltration was classified as chronic inflammation (here in short inflammation). Proportions of prostates with inflammation in CMV seronegative (IgG-; n=17) and CMV seropositive (IgG+; n=24) post-mortem donors were compared with Fisher's exact test.

**B)** An association between CMV-pp71 epithelial abundance and presence of inflammation in CMV seronegative (IgG-) and CMV seropositive (IgG+) post-mortem donors was examined with Kruskal-Wallis multiple comparisons test. Ns is non-significant. Graph shows all datapoints and mean±SD.

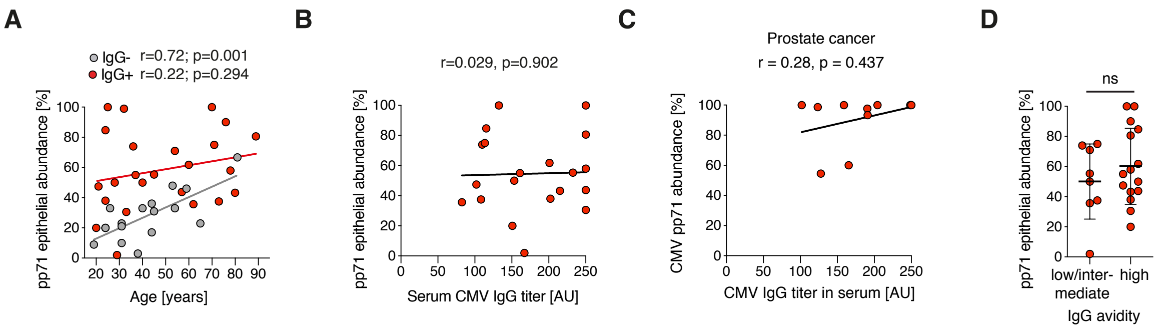


**Supplemental Figure 2: CMV abundance and associations with age, CMV IgG titer and CMV IgG avidity**

**A)** Pearson correlation between CMV-pp71 epithelial abundance and age in CMV seronegative (IgG-; n=17) and CMV seropositive (IgG+; n=24) post-mortem donors separately. **B)** Pearson correlation analysis of CMV-pp71 epithelial abundance in the post-mortem prostate donors and serum CMV IgG titer in seropositive subjects. AU = arbitrary units. Samples that had titers of 250 or higher were considered to have the titer 250 AU in the analysis. **C)** Graph showing that CMV abundance in cancer (% CMV pp71+ cancer areas) is not associated with CMV IgG titer in serum of CMV seropositive patients (n=10). A Spearman correlation analysis was performed. AU = arbitrary units. **D)** CMV-pp71 epithelial abundance (mean ±SD) in CMV seropositive post-mortem donors (n=23) with low or intermediate serum IgG avidity were compared with donors with high serum IgG avidity with two-sided un-paired t-test. Graph shows all datapoints and mean±SD.

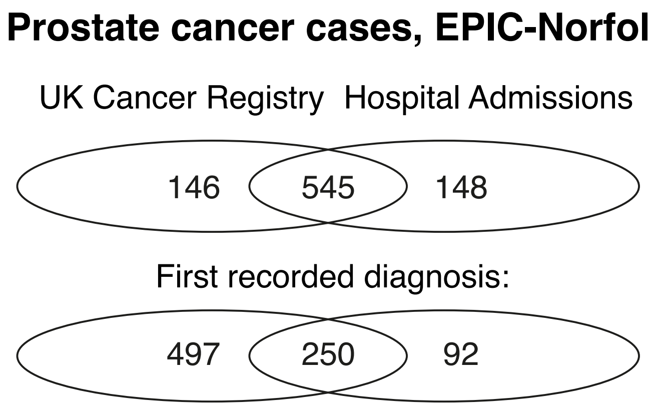


**Supplemental Figure 3: Prostate cancer cases in EPIC-Norfolk**

Cases were registered in the UK Cancer Registry until 2016 and from hospital admissions until 2018. The venn diagram show overlap in total cases registered and when the diagnosis was first recorded. The cases include prevalent (diagnosed prior to CMV IgG determination) and incident (diagnosed after CMV IgG determination) prostate cancer patients.

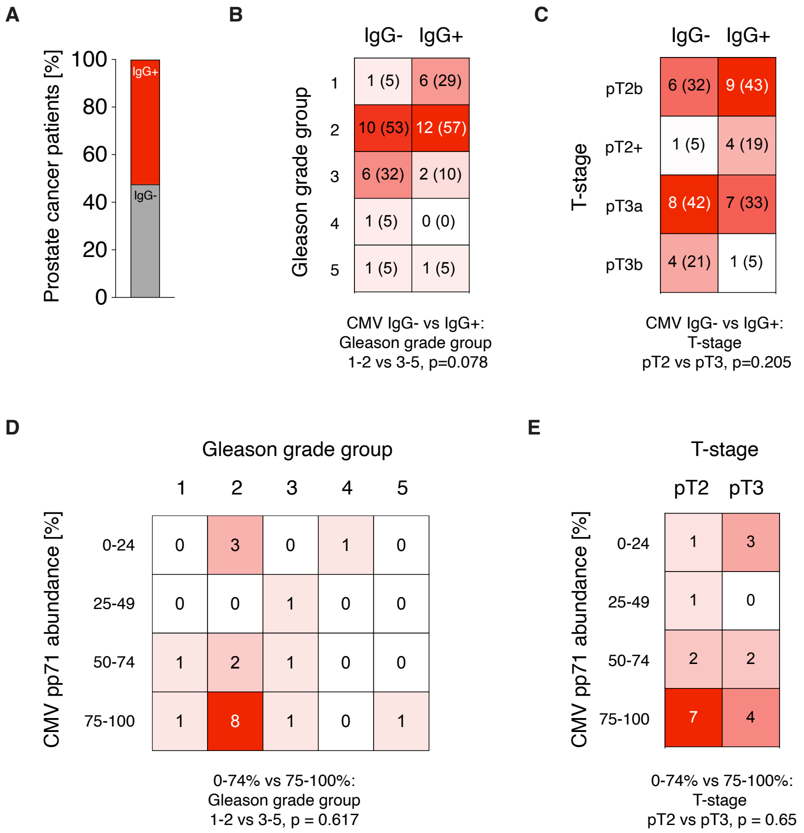


**Supplemental Figure 4: CMV is not associated with Gleason grade group or T-stage in the prostatectomy cohort**

**A)** Percentage of prostate cancer patients aged 50-68 years (n=40) that were CMV seronegative (IgG-) or CMV seropositive (IgG+). Data is presented as a stacked bar graph with percentage of samples. **B-C)** Heat maps showing Gleason grade group (B) and pT-stage (C) comparing frequency of Gleason grade 1-2 vs 3-5 and pT-stage pT2 vs pT3 in CMV IgG- and CMV IgG+ prostate cancer patients with Fisher's exact test. Number of samples are shown in heat maps (n (%). **D-E)** Heat map showing Gleason grade group (D) and pathological T-stage (E) and different abundance of CMV in primary prostate cancer (n=20). Frequency of Gleason grade group (1-2 vs 3-5) and T-stage (pT2 vs pT3) was examined with Fisher's exact test comparing tumors with 1-74% compared to 75-100% CMV abundance. Number of samples are shown in the heat maps. pT-stage = pathological T-stage. T2+ = organ-confined disease with positive surgical margin on prostatectomy sample. The cancer does not grow outside the prostate capsule. pT2b = greater than one-half of one prostate lobe with cancer. pT3a = growth outside of the prostate gland. pT3b = Invasion into seminal vesicle(s)

| | CMV IgG$^{-}$ (n=3254) | CMV IgG$^{+}$ (n=4401) | p-value |
|---|---|---|---|
| CMV IgG serostatus in % | 43% | 57% | |
| Age, mean years (SD) | 58.6 (9.1) | 61.5 (9.2) | <0.001 |
| Smoking | 62.9% | 68.2% | 0.004 |
| Alcohol usage, mean (SD) | 10.8 (12.3) | 10.0 (11.2) | 0.570 |
| Townsend index, mean (SD) | -2.20 (2.0) | -2.05 (2.2) | 0.004 |
| Education level | 64.3% | 59.5% | 0.030 |
| Social class | 60.2% | 58.3% | 0.098 |
| Marital Status | 87.0% | 88.6% | 0.017 |
| BMI ≥ 30 | 11.6% | 13.7% | 0.023 |
| Waist-hip ratio ≥ 0.9 | 67.9% | 71.5% | 0.241 |
| Diabetes Mellitus | 2.4% | 3.2% | 0.437 |
| Ethnicity Caucasian | 99.9 | 99.4 | 0.001 |

**Supplemental Table 1: Characteristics of the male CMV EPIC-Norfolk cohort**

Analyses were adjusted for age and were performed with logistic or linear regression. SD = Standard Deviation, CMV = Human Cytomegalovirus, IgG = Immunoglobulin G, Smoking: ever smokers, Alcohol usage: units per week, Education level: A-level or higher, Social class: skilled non-manual worker or higher, BMI=Body Mass Index, Ethnicity: Caucasian or other than Caucasian. IgG$^{-}$ = seronegative; IgG$^{+}$ = seropositive.

| | 1st Health Check (n=5894) | | 2nd Health Check (n=3846) | |
|---|---|---|---|---|
| **Age (y)** | **CMV IgG$^-$** | **CMV IgG$^+$ (%)** | **CMV IgG$^-$** | **CMV IgG$^+$ (%)** |
| 40-49 | 562 (55.4) | 453 (44.6) | 120 (56.9) | 91 (43.1) |
| 50-59 | 914 (46.0) | 1071 (54.0) | 605 (49.8) | 610 (50.2) |
| 60-69 | 741 (39.7) | 1127 (60.3) | 582 (42.5) | 788 (57.5) |
| 70-79 | 326 (31.8) | 700 (68.2) | 368 (36) | 655 (64.0) |
| 80-89 | 0 (0) | 0 (0) | 8 (29.6) | 19 (70.4) |

**Supplemental Table 2: Prevalence of CMV seropositivity increase with age in men**

Blood was drawn in 1st health check (1993-1998) and 2nd health check (1998-2000) from male study participants in EPIC-Norfolk. For 1160 men, blood was analyzed for CMV serostatus at both 1st and 2nd health check. IgG$^-$ = seronegative; IgG$^+$ = seropositive.

| | **Person-time (years)** | **Crude incidence rate ratio (CI 95%)** | **p-value** |
|---|---|---|---|
| CMV IgG- | 61705.1 | | |
| CMV IgG+ | 76946.9 | 1.14 (0.99-1.32) | 0.068 |

**Supplemental Table 3: Crude incidence rate ratios for prostate cancer incidence**
Prostate cancer patients are defined as incident cases, i.e. that their diagnoses are registered after CMV serostatus examination. CMV seropositive men are compared to CMV seronegative patients. Person-time in years and crude incidence rate ratios were calculated. Two-sided tests were calculated to determine a p-value.

| **Incident Prostate cancer (n=278)** | **CMV IgG$^-$** | **CMV IgG$^+$** | **p-value** |
|---|---|---|---|
| Local prostate cancer (%) | 74 (69) | 125 (73) | |
| Advanced prostate cancer (%) | 33 (31) | 46 (27) | 0.487 |

**Supplemental Table 4: CMV serostatus is not associated with advanced prostate cancer at diagnosis**

The statistical analysis was made by age-adjusted logistic regression. Data on cancer stage was extracted from the UK Cancer Registry. Local and advanced prostate cancer are defined accordingly: localized prostate cancer: local growth in prostate; T2 or lower, N0, M0. Advanced prostate cancer: tumor has invaded surrounding tissue and/or presence of lymph node metastases and/or presence of distant metastases; T3 or higher, or N1 or M1. IgG$^-$ = seronegative; IgG$^+$ = seropositive.

| | Person-time (years) | Crude incidence rate ratio (CI 95%) | p-value |
|---|---|---|---|
| **All-cause mortality in patients with prostate cancer** | | | |
| CMV IgG- | 1919.5 | | |
| CMV IgG+ | 2719.9 | 1.33 (1.04-1.71) | 0.020 |
| **Prostate cancer mortality in patients with prostate cancer** | | | |
| CMV IgG- | 1919.5 | | |
| CMV IgG+ | 2719.9 | 1.44 (0.99-2.13) | 0.046 |
| **All-cause mortality in all men** | | | |
| CMV IgG- | 60733.6 | | |
| CMV IgG+ | 76594.5 | 1.41 (1.31-1.52) | <0.001 |
| **Prostate cancer mortality in all men** | | | |
| CMV IgG- | 60733.6 | | |
| CMV IgG+ | 76594.5 | 1.56 (1.16-2.13) | 0.003 |

**Supplemental Table 5: Crude incidence rate ratios for prostate cancer mortality and all-cause mortality**

Prostate cancer patients are defined as incident cases, i.e. that their diagnoses are registered after CMV serostatus examination. CMV seropositive patients are compared to CMV seronegative patients. Person-time in years and crude incidence rate ratios were calculated. Two-sided tests were calculated to determine a p-value.
